# Age as a moderator of a brief alcohol intervention among injury patients in Northern Tanzania

**DOI:** 10.64898/2026.06.10.26355118

**Authors:** Winfrida C. Mwita, João Ricardo Nickenig Vissoci, Joao Vitor Perez de Souza, Ashley J. Phillips, Baraka Moshi, Kim Madundo, Florida J. Muro, Blandina T. Mmbaga, Sia E. Msuya, Catherine A. Staton

## Abstract

**Background:** Alcohol use is a leading modifiable risk factor for injury in sub-Saharan Africa. In Tanzania, young people (≤24 years) experience greater alcohol-related harm despite drinking less frequently than adults. Punguza Pombe kwa Afya Yako (PPKAY) is a culturally adapted, brief intervention for injury patients in Tanzania. This study examined whether age moderates its effectiveness.

**Methods:** We conducted an exploratory secondary analysis of baseline and 3-month data from the PPKAY randomized trial among injury patients aged ≥18 years at Kilimanjaro Christian Medical Centre, Tanzania. Eligible participants reporting alcohol use before injury, AUDIT ≥8, or positive breathalyzer were randomized to usual care or PPKAY with SMS boosters. The primary outcome was binge drinking days. Count outcomes were analyzed using negative binomial regression with robust SEs and continuous outcomes using mixed-effects models. Effect modification was assessed using a three-way interaction (Time × intervention × Age).

**Results:** Among 543 participants (mean age 36.8 years; 16.2% aged 18–24), age moderated the intervention effect for drinking days (IRR = 0.27, 95% CI 0.07–0.98; p = 0.046) and drinks consumed (IRR = 0.17, 95% CI 0.04–0.77; p = 0.021). The intervention reduced 4 drinking days (95% CI −7.1 to −0.8) and 27.5 drinks (95% CI −42.8 to −12.2) among young people, while adults showed reductions in both arms, without intervention-specific effect.

**Conclusion:** The effects of ED-based brief alcohol interventions are not uniform, varying across both age groups and alcohol-related outcomes. We found a greater responsiveness in drinking frequency and quantity reported among young people.

## Introduction

Alcohol use is a major preventable contributor to global morbidity and mortality, accounting for over 2.6 million deaths annually and 4.7% of the global burden of disease (1) . The burden is disproportionately high in low- and middle-income countries (LMICs) (1), where health systems face substantial constraints in managing alcohol-related harm (2). The WHO African Region reports among the highest alcohol-attributable death rates worldwide (1) . In Tanzania, per capita alcohol consumption stands at 9.4 liters, exceeding the regional average of 6.4 liters (1). Alcohol use is relatively higher in the Kilimanjaro region (3), where nearly 30% of injury patients presenting to the Kilimanjaro Christian Medical Centre (KCMC) emergency department test positive for alcohol on arrival (4), underscoring the need for alcohol harm reduction strategies within trauma care.

Brief interventions (BIs), including single-session approaches grounded in motivational interviewing (MI), have been shown to reduce alcohol consumption and related harm, particularly in primary care and emergency department settings (5–7). Rooted in the Screening, Brief Intervention, and Referral to Treatment (SBIRT) model, BIs aim to enhance readiness to change through empathetic, non-confrontational counselling (8,9). BIs are recommended by the World Health Organization as a cost-effective health system response to harmful alcohol use (10). However, findings on BI effectiveness have not been consistent across studies. Pragmatic trials in ED settings have reported limited or no additional benefit of brief interventions (11), with such null findings attributed, in part, to reduced intervention fidelity in real-world settings (12). Implementation in busy ED settings may be affected by competing clinical priorities and limited time (13), which may further compromise intervention delivery. In addition to these system-level challenges, prior studies have suggested that overall intervention effects may obscure important subgroup differences, with evidence of heterogeneity and inconsistent findings across populations, particularly by sex and other patient characteristics (7,14,15). Consistent with this, patient-level characteristics such as baseline drinking severity (16), alcohol attribution (17,18), and age (18,19) have been identified as potential moderators of BI outcomes. Evidence from LMICs, particularly among trauma patients, remains limited (6).

In response to the high burden of alcohol-related injuries, the Punguza Pombe kwa Afya Yako (PPKAY) intervention, translated as “*Reduce Alcohol for Your Health*” was developed and culturally adapted for use in Northern Tanzania. PPKAY is a 30-minute, nurse-delivered, motivational interviewing-based brief intervention tailored to the Tanzanian ED settings (20). Feasibility and acceptability of PPKAY delivery by non-specialist providers were demonstrated (21); a subsequent pragmatic randomised trial found that patients who received the PPKAY intervention (pooled PPKAY + standard and personalised booster) showed significant reductions in binge drinking days, total alcohol consumption, and drinking frequency at 3-month follow-up, compared to usual care (22). Building on these findings, the present study examines potential heterogeneity in treatment response by evaluating whether age moderates the effectiveness of the PPKAY intervention.

Age is increasingly recognised as a potential moderator of brief intervention efficacy (18,19,23). Behavioural change models such as the Transtheoretical Model suggest that motivation to change may vary across the life course (24). Younger individuals, particularly those aged 18 to 24, are more likely to engage in binge drinking and impulsive alcohol use yet may also respond differently to interactive or tailored interventions (25). Studies from high-income countries have shown mixed results: some suggest stronger BI effects among younger people, while others report better outcomes in older adults with more entrenched drinking patterns (17–19,23). A meta-analysis by Elzerbi et al. noted that brief interventions were less effective among ED patients with injuries compared to non-injured patients and highlighted the potential for patient characteristics, including age, to contribute to the variability in response (26). However, evidence regarding age-related differences in BI responsiveness within LMIC emergency care settings remains scarce.

Building on previous work demonstrating significant differences in alcohol use patterns and consequences between young people (18–24 years) and adults (25+ years) among injury patients at KCMC (27), the present study examines whether age moderates the effectiveness of the PPKAY intervention on alcohol-related outcomes. Although age groups differ in baseline drinking patterns, it remains unclear whether these differences translate into differential responsiveness to brief intervention. Understanding the moderating role of age has important implications for optimising intervention delivery within trauma care settings.

## Methods

### Study Design and Setting

This study is a secondary analysis from the pragmatic randomized adaptive clinical trial (PRACT) (ClinicalTrials.gov: identifier: NCT04535011), which enrolled participants to evaluate the effectiveness of a culturally adapted brief intervention, PPKAY, to reduce harmful and hazardous alcohol use among injury patients in Tanzania (28). Data for this analysis were collected between October 2020 and March 2025. The trial was conducted at the ED of KCMC, a tertiary referral hospital located in Moshi, Northern Tanzania. For this analysis, we used data from the parent trial from a parallel, single-blind, block-randomised design with allocation to usual care or PPKAY (delivered either alone or with a standard or personalised SMS booster). For this analysis, following the analytical protocol of PRACT’s stage I, intervention groups were pooled and compared to usual care because the core brief intervention content was identical across arms, thereby increasing statistical power to evaluate effect modification by age. Detailed randomisation, allocation concealment, and implementation procedures are described in the published PRACT protocol (28).

### Participants

Eligible participants were injury patients (≥18 years) who presented to the KCMC ED seeking care for an acute injury. Inclusion criteria required that patients were not clinically intoxicated and met at least one of the following: AUDIT score of ≥8, positive breathalyser test result (>0.0 g/dL), or self-reported alcohol use prior to the injury. Patients were excluded if they were too ill to participate, unable to communicate, intoxicated at the time of consent, or declined informed consent. Additional exclusion criteria included non-Swahili speakers, lack of access to a mobile phone for follow-up text messaging, or not having resided in East Africa for at least five years. Written informed consent was obtained from all participants prior to enrolment.

### Intervention

Participants randomised to the intervention arm received a single-session, nurse-delivered, 15-minute PPKAY brief intervention grounded in motivational interviewing principles, addressing alcohol use patterns, perceived consequences, readiness to change, and collaborative goal-setting. Those in the PPKAY + Standard Booster group received weekly SMS for one year containing general motivational messages. Those in the PPKAY + The personalised booster group received customised SMS messages aligned with the participant’s individual goals. Usual care participants did not receive any alcohol-specific counselling or messages.

## Measures

### Primary outcome

The primary outcome was the number of binge drinking days in the past 28 days, assessed at both baseline and 3-month follow-up. Participants were asked, “During the last four weeks, how many days did you have 5 or more drinks on one occasion (for men) or 4 or more drinks on one occasion (for women)?”

### Secondary outcomes

Secondary outcomes included alcohol-related consequences measured using the Drinker Inventory of Consequences (DrInC) (29), number of standard drinks consumed, number of drinking days, Alcohol Use Disorders Identification Test (AUDIT) score (30) , and depressive symptoms measured using the Patient Health Questionnaire-9 (PHQ-9) (31), assessed at both baseline and 3-month follow-up. The DrInC and AUDIT questions referred to a three-month time window; number of drinking days and number of standard drinks consumed referred to the previous four weeks; and PHQ-9 referred to the previous two weeks. All outcomes were self-reported and collected during standard follow-up windows using a timeline follow-back procedure (32).

### Moderator variable

The moderator of interest was age, categorized as 18–24 years and ≥25 years. This categorization reflects public health frameworks recognizing late adolescence and early adulthood as high-risk periods for alcohol use and injury and aligns with prior evidence of age-related differences in alcohol harm in this setting.

### Statistical analysis

All analyses were conducted using Stata version 15.0 (StataCorp, College Station, TX) (32). Descriptive statistics were calculated overall and stratified by age group. Baseline descriptive variables included age, sex, tribe, education level, employment status and intervention group allocation. Baseline comparisons were performed using t-tests or Wilcoxon rank-sum tests for continuous variables and chi-square tests for categorical variables. Analyses followed an intention-to-treat principle using the available follow-up data and were restricted to participants with both baseline and 3-month follow-up measurements.

To assess whether age moderated the effectiveness of PPKAY over time, we used regression models included fixed effects for time (baseline vs. 3 months), intervention group (PPKAY vs. usual care), age group (18–24 vs. ≥25 years), and a three-way interaction term (Time × Intervention × Age Group). The primary outcome (binge drinking days) and other count outcomes (drinking days and number of drinks consumed) were analyzed using negative binomial regression with cluster-robust standard errors at the participant level. Secondary outcomes (AUDIT, PHQ-9, and DrInC scores) were log-transformed using ln (score + 1) and analysed using linear mixed-effects models with random intercepts for participants. Count models and DrInC models were adjusted for sex, baseline AUDIT score, and baseline PHQ-9 score to account for baseline alcohol use severity and depressive symptoms that may influence outcomes over time. The AUDIT model was adjusted for sex and baseline PHQ-9 only, and the PHQ-9 model for sex and baseline AUDIT only to avoid adjusting for baseline values of the same construct. Model assumptions for continuous outcomes were assessed by inspecting residual and Q–Q plots for homoscedasticity and normality, and overdispersion for count outcomes was assessed by comparing Poisson and negative binomial model fit. In age-stratified models, predicted means were estimated at baseline and 3 months using marginal predictions, and differences in predicted means were used to derive within-arm changes and difference-in-differences estimates. Results are reported as incidence rate ratios (IRRs) for count outcomes and β coefficients for continuous outcomes with 95% confidence intervals.

Statistical significance was set at p < 0.05 (two-sided). Because the parent trial was powered to detect overall intervention effects rather than effect modification, moderation analyses were considered exploratory, and interaction estimates were interpreted in light of confidence intervals. Sensitivity analyses included repeating models using multiple imputation by chained equations (20 imputations) under the assumption of missing at random and re-estimating moderation models with age entered as a continuous, mean-centred variable.

### Ethical approval

The parent PRACT trial was approved by institutional review boards in Tanzania and the United States (Duke University IRB Pro000103724; Kilimanjaro Christian Medical University College Ethics Committee Certificate #2457; Tanzanian National Institute for Medical Research NIMR/HQ/R.8a/Vol.IX/3425) and was prospectively registered on <u>ClinicalTrials.gov</u> (identifier: NCT04535011). All participants provided written informed consent prior to enrolment. This secondary analysis received additional ethical approval from the Kilimanjaro Christian Medical University College Ethics Committee (Certificate #2708) and the Tanzanian National Institute for Medical Research (NIMR/HQ/R.8a/Vol.IX/4839). Data were de-identified prior to analysis, and no additional consent was required.

## Results

### Characteristics of the study participants

At baseline, the mean age of participants was 36.6 years (SD 12.7), with 83.8% aged ≥25 years. Most participants were male (93.5%) and of Chagga ethnicity (57.5%), and the majority were employed (96.4%) (Table 1).

**Table 1.** Baseline socio-demographic characteristics of the study participants, stratified by age groups (N=543)

|  |  | 18–24 years, n (%) | ≥25 years, n (%) |
| --- | --- | --- | --- |
| Variable | Total, n (%) | (n = 88) | (n = 455) |
| Age, years |  |  |  |
| Mean (SD) | 36.6 (12.7) | 22.3 (1.6) | 39.3 (12.0) |
| Median (Q1–Q3) | 34 (27–44) | 23 (21–24) | 36 (30–46) |
| Sex |  |  |  |
| Male | 508 (93.5) | 83 (94.3) | 425 (93.4) |
| Female | 35 (6.5) | 5 (5.7) | 30 (6.6) |
| Tribe |  |  |  |
| Chagga | 312 (57.5) | 54 (61.4) | 258 (56.7) |
| Pare | 64 (11.8) | 6 (6.8) | 58 (12.7) |
| Other | 167 (30.7) | 28 (31.8) | 139 (30.6) |
| Education level |  |  |  |
| No formal education | 7 (1.4) | 0 (0.0) | 7 (1.7) |
| Primary | 276 (56.6) | 37 (51.4) | 239 (57.5) |
| Secondary | 92 (18.8) | 23 (31.9) | 69 (16.6) |
| Vocational & Higher | 113 (23.2) | 12 (16.7) | 101 (24.3) |
| Employment |  |  |  |
| Student | 9 (1.7) | 7 (8.0) | 2 (0.4) |
| Unemployed | 10 (1.9) | 0 (0.0) | 10 (2.2) |
| Employed | 522 (96.4) | 81 (92.0) | 441 (97.4) |

### Primary and Secondary Outcomes

At baseline, adults reported slightly higher drinking frequency and quantity than younger participants, with median drinking days of 5 compared with 4 and median drinks of 22.4 compared with 16.0 in the past 28 days. By 3 months, alcohol use declined substantially in both age groups, with median binge-drinking days, drinking days, and number of drinks all decreasing to zero. AUDIT and DrInC scores also declined in both age groups, while PHQ-9 scores remained low over time (Table 2).

**Table 2.**
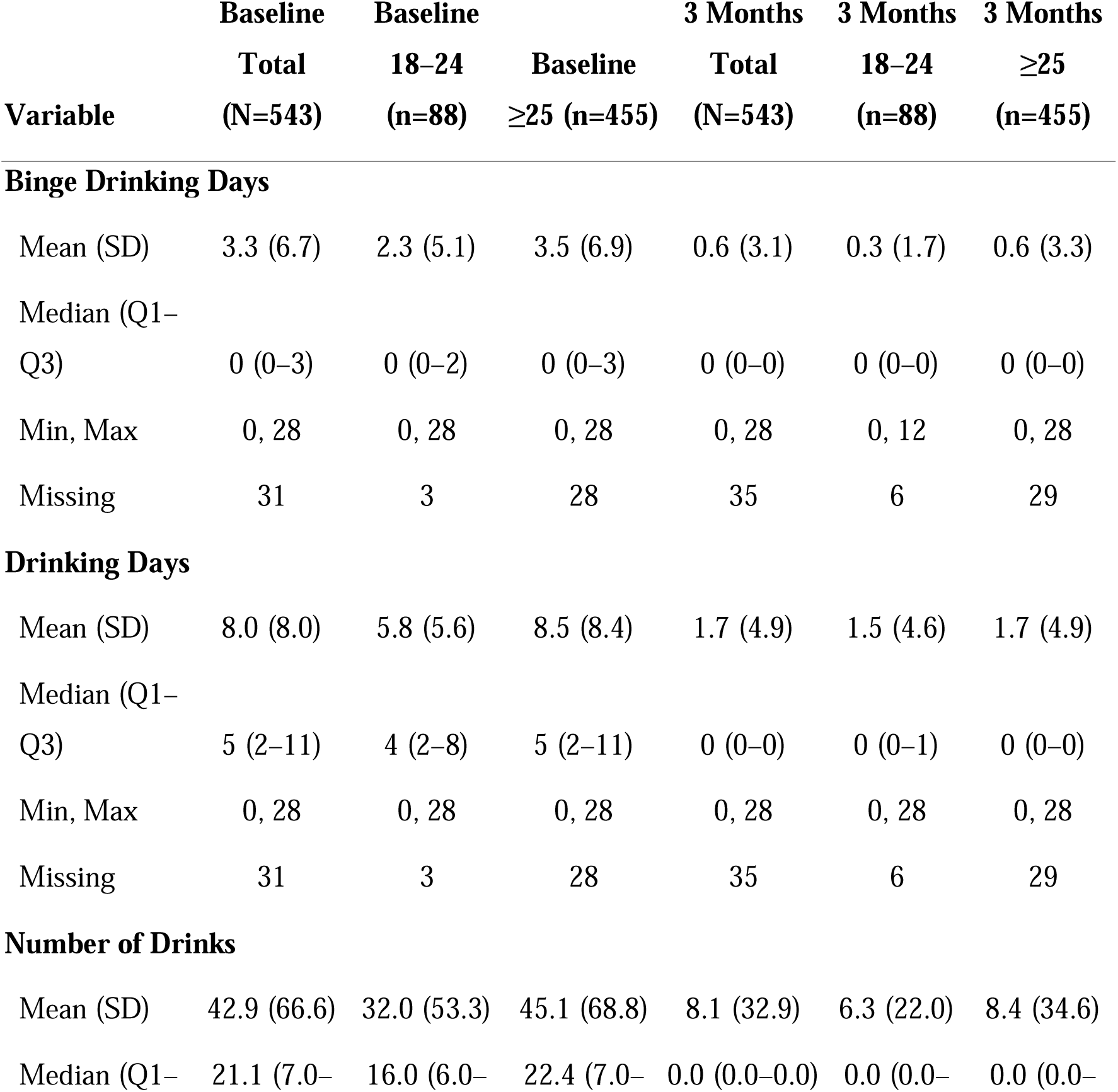
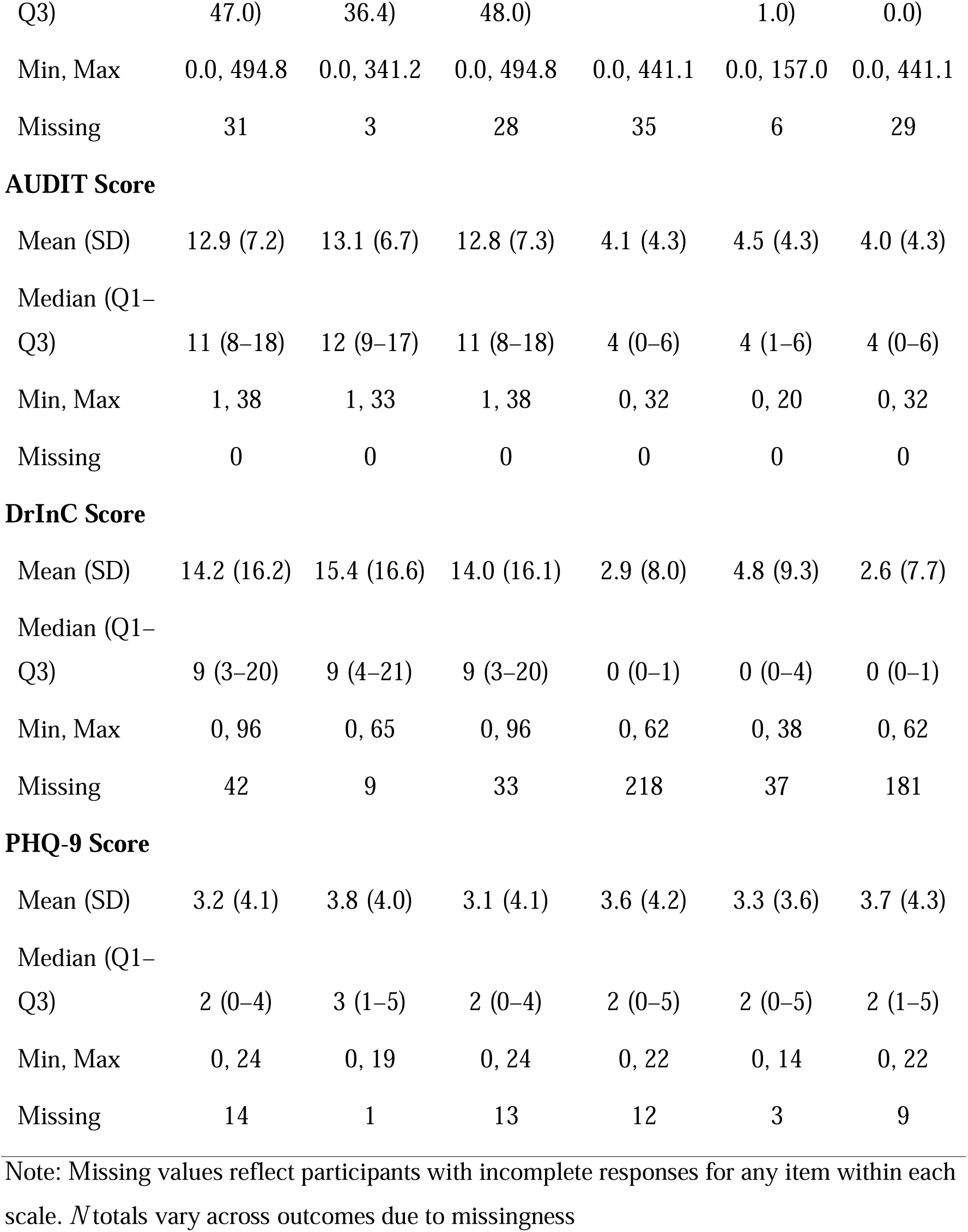
Primary and secondary outcomes by age group at baseline and 3 months (N=543)

| Variable | Baseline<br>Total<br>(N=543) | Baseline<br>18–24<br>(n=88) | Baseline<br>≥25 (n=455) | 3 Months<br>Total<br>(N=543) | 3 Months<br>18–24<br>(n=88) | 3 Months<br>≥25<br>(n=455) |
| --- | --- | --- | --- | --- | --- | --- |
| **Binge Drinking Days** |  |  |  |  |  |  |
| Mean (SD) | 3.3 (6.7) | 2.3 (5.1) | 3.5 (6.9) | 0.6 (3.1) | 0.3 (1.7) | 0.6 (3.3) |
| Median (Q1–<br>Q3) | 0 (0–3) | 0 (0–2) | 0 (0–3) | 0 (0–0) | 0 (0–0) | 0 (0–0) |
| Min, Max | 0, 28 | 0, 28 | 0, 28 | 0, 28 | 0, 12 | 0, 28 |
| Missing | 31 | 3 | 28 | 35 | 6 | 29 |
| **Drinking Days** |  |  |  |  |  |  |
| Mean (SD) | 8.0 (8.0) | 5.8 (5.6) | 8.5 (8.4) | 1.7 (4.9) | 1.5 (4.6) | 1.7 (4.9) |
| Median (Q1–<br>Q3) | 5 (2–11) | 4 (2–8) | 5 (2–11) | 0 (0–0) | 0 (0–1) | 0 (0–0) |
| Min, Max | 0, 28 | 0, 28 | 0, 28 | 0, 28 | 0, 28 | 0, 28 |
| Missing | 31 | 3 | 28 | 35 | 6 | 29 |
| **Number of Drinks** |  |  |  |  |  |  |
| Mean (SD) | 42.9 (66.6) | 32.0 (53.3) | 45.1 (68.8) | 8.1 (32.9) | 6.3 (22.0) | 8.4 (34.6) |
| Median (Q1– | 21.1 (7.0– | 16.0 (6.0– | 22.4 (7.0– | 0.0 (0.0–0.0) | 0.0 (0.0– | 0.0 (0.0– |
| Q3) | 47.0) | 36.4) | 48.0) |  | 1.0) | 0.0) |
| Min, Max | 0.0, 494.8 | 0.0, 341.2 | 0.0, 494.8 | 0.0, 441.1 | 0.0, 157.0 | 0.0, 441.1 |
| Missing | 31 | 3 | 28 | 35 | 6 | 29 |
| **AUDIT Score** |  |  |  |  |  |  |
| Mean (SD) | 12.9 (7.2) | 13.1 (6.7) | 12.8 (7.3) | 4.1 (4.3) | 4.5 (4.3) | 4.0 (4.3) |
| Median (Q1–<br>Q3) | 11 (8–18) | 12 (9–17) | 11 (8–18) | 4 (0–6) | 4 (1–6) | 4 (0–6) |
| Min, Max | 1, 38 | 1, 33 | 1, 38 | 0, 32 | 0, 20 | 0, 32 |
| Missing | 0 | 0 | 0 | 0 | 0 | 0 |
| **DrInC Score** |  |  |  |  |  |  |
| Mean (SD) | 14.2 (16.2) | 15.4 (16.6) | 14.0 (16.1) | 2.9 (8.0) | 4.8 (9.3) | 2.6 (7.7) |
| Median (Q1–<br>Q3) | 9 (3–20) | 9 (4–21) | 9 (3–20) | 0 (0–1) | 0 (0–4) | 0 (0–1) |
| Min, Max | 0, 96 | 0, 65 | 0, 96 | 0, 62 | 0, 38 | 0, 62 |
| Missing | 42 | 9 | 33 | 218 | 37 | 181 |
| **PHQ-9 Score** |  |  |  |  |  |  |
| Mean (SD) | 3.2 (4.1) | 3.8 (4.0) | 3.1 (4.1) | 3.6 (4.2) | 3.3 (3.6) | 3.7 (4.3) |
| Median (Q1–<br>Q3) | 2 (0–4) | 3 (1–5) | 2 (0–4) | 2 (0–5) | 2 (0–5) | 2 (1–5) |
| Min, Max | 0, 24 | 0, 19 | 0, 24 | 0, 22 | 0, 14 | 0, 22 |
| Missing | 14 | 1 | 13 | 12 | 3 | 9 |

Note: Missing values reflect participants with incomplete responses for any item within each scale. *N* totals vary across outcomes due to missingness

### Moderating effect of age on intervention outcomes

Predicted average binge-drinking days decreased from baseline to three months in both age groups, with similar downward trends across the intervention and usual-care arms among young people (18–24 years) and adults (≥25 years) (Figure 2A–B). The three-way interaction between time, intervention, and age group showed a wider confidence interval, indicating both groups had reductions within similar ranges.

**Figure 1:**
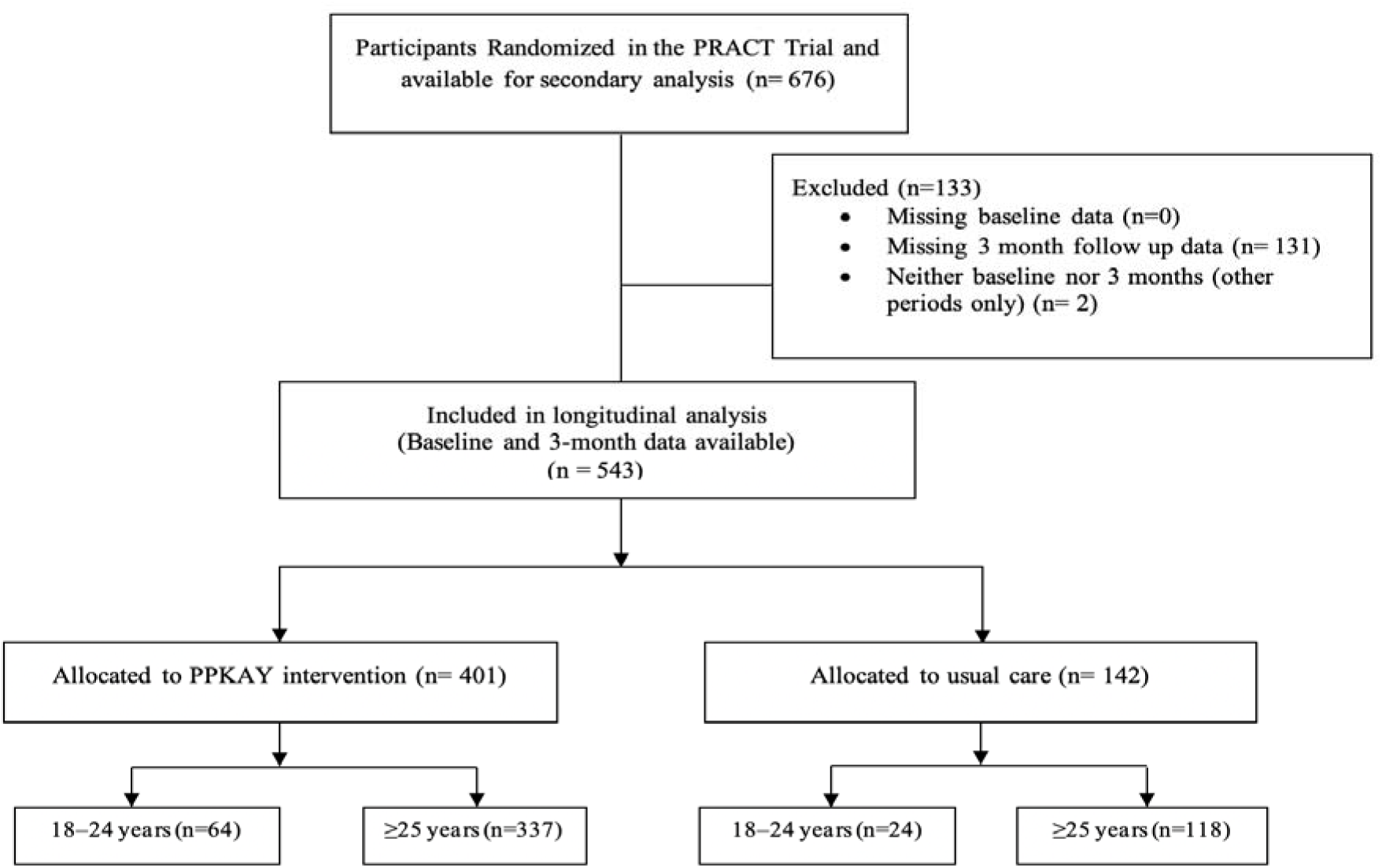
Participant flow chart for the secondary longitudinal analysis of the PRACT trial

**Figure 2.**
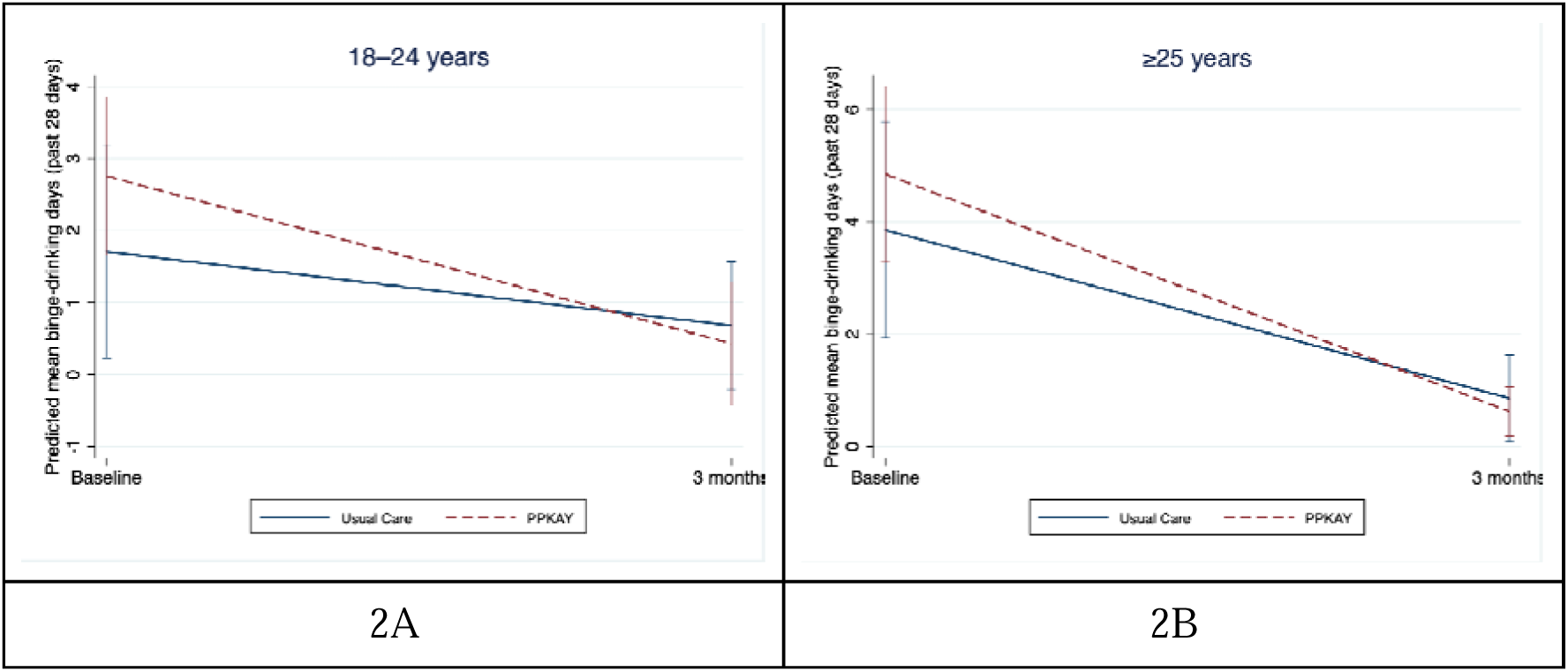
Predicted binge-drinking days at 3 months by intervention arm, stratified by age group: **A**. Young people (18-24 years); **B**. Adults (≥25 years)

However, moderation by age group was observed for drinking days and number of drinks consumed. The intervention effect showed confidence interval ranges endorsed wide but consistent differences between groups for drinking days (IRR = 0.27, 95% CI 0.07–0.98) and number of drinks (IRR = 0.17, 95% CI 0.04–0.77). No moderation effects were observed for AUDIT, DrInC, or PHQ-9 scores (Table 3).

**Table 3:**
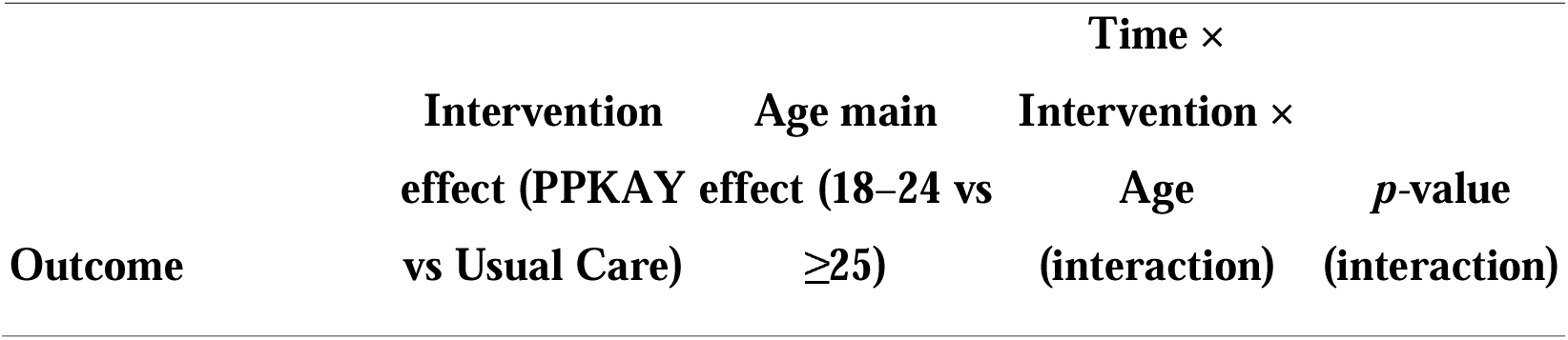
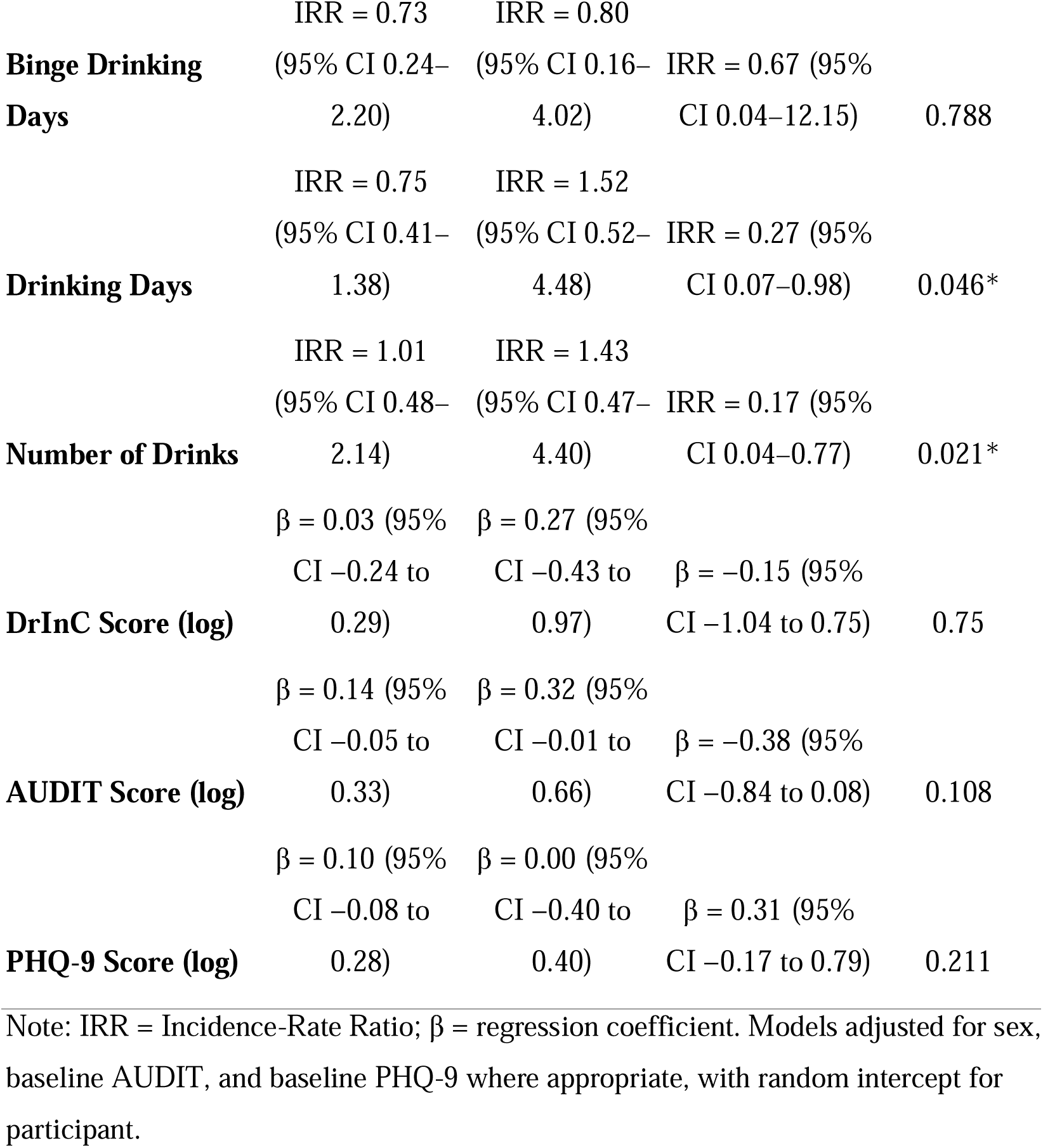
Moderating effect of age on intervention outcomes at 3 months.

### Age-stratified intervention effect on drinking quantity and frequency

Among participants aged 18–24 years, reductions in alcohol consumption were observed in both the intervention and usual-care groups at the 3-month follow-up. However, the intervention was associated with an additional reduction of 4.0 drinking days (95% CI −7.1 to −0.8) and 27.5 drinks (95% CI −42.8 to −12.2) over the 28-day period compared with usual care (Figures 3–4; Table S1). The intervention was not associated with a significant additional reduction in binge-drinking days in this age group (DiD −1.4 days, 95% CI −3.3 to 0.6) (Table S1).

**Figure 3.**
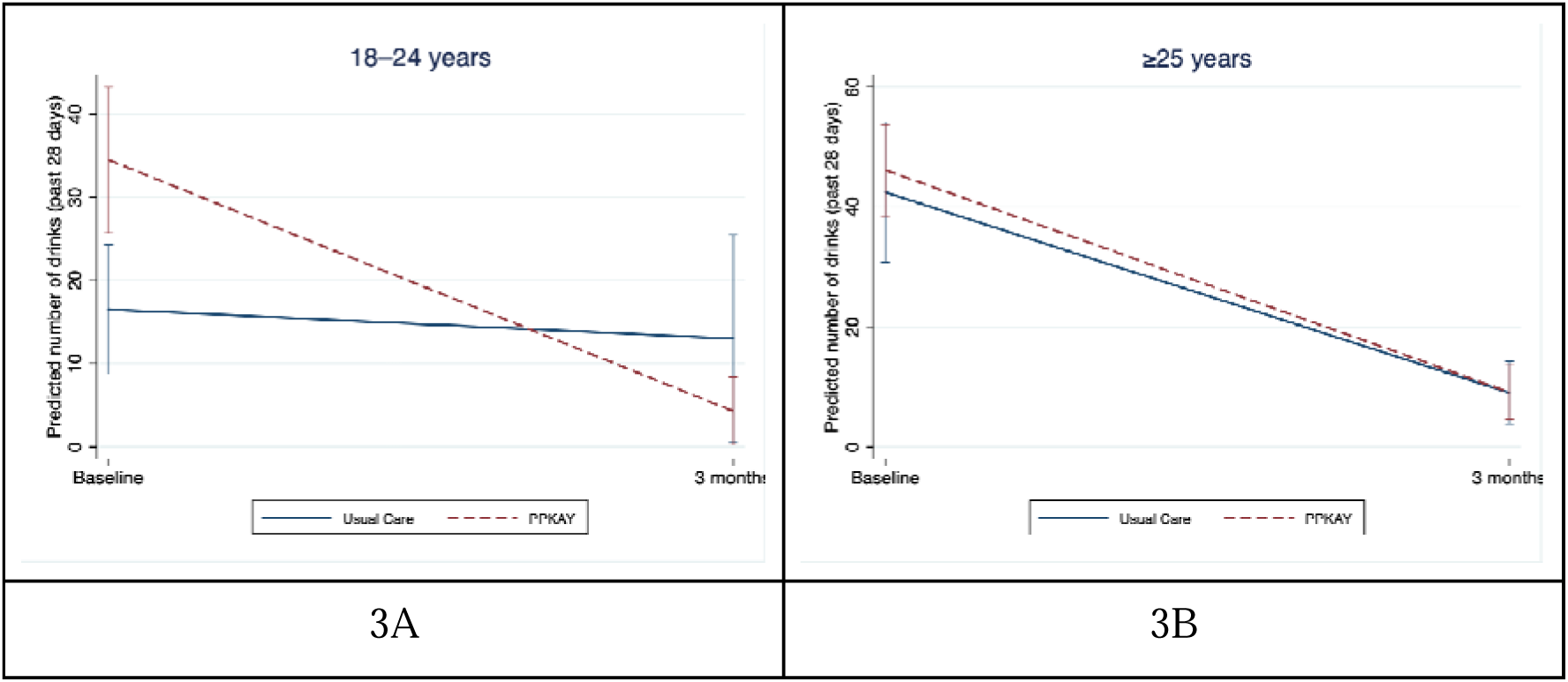
Predicted Number of Drinks (Past 28 Days) at 3 Months by Intervention Arm, Stratified by Age Group: **A**. Young people (18-24 years); **B**. Adults (≥25 years)

**Figure 4.**
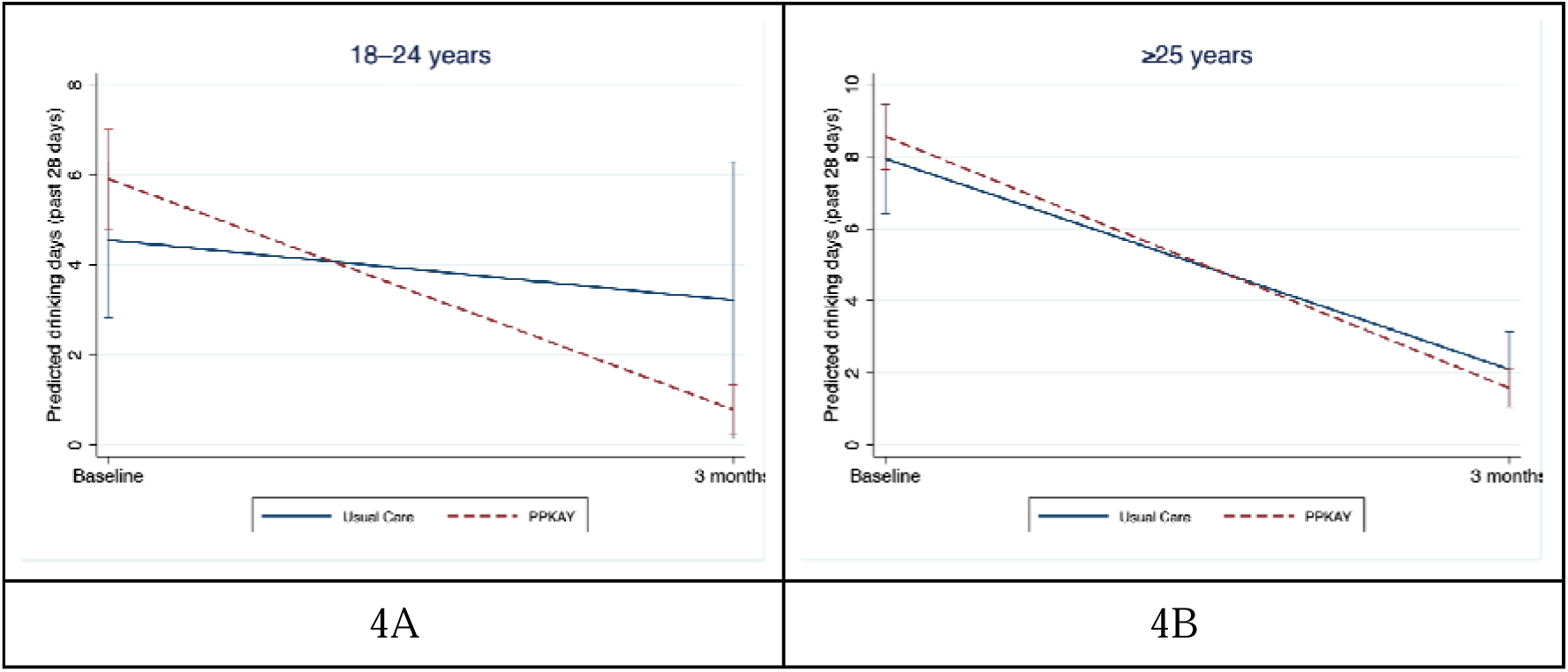
Predicted Number of Drinking Days (Past 28 Days) at 3 Months by Intervention Arm, Stratified by Age Group: **A**. Young people (18-24 years); **B**. Adults (≥25 years)

Among adults aged ≥25 years, alcohol consumption declined from baseline to three months in both the intervention and usual-care groups, with no evidence of an intervention-specific effect for drinking days (DiD −1.1 days, 95% CI −3.2 to 0.9), number of drinks (DiD −3.2 drinks, 95% CI −17.1 to 10.8), or binge-drinking days (DiD −1.3 days, 95% CI −3.8 to 1.2) (Figures 3–4; Table S1).

### Sensitivity Analysis

Multiple imputation analyses produced estimates similar to those from the complete-case models. In the imputed models, the time × intervention × age interaction showed similar confidence intervals for the number of drinks outcome and drinking days, while interactions for binge drinking days, AUDIT, DrInC, and PHQ-9 scores were not observed (Table S3). When age was modelled as a continuous variable (per one-year increase in age), the time × intervention × age interaction terms were not observed across outcomes, indicating that the observed age moderation effect for drinking days and number of drinks did not follow a linear trend with increasing age (Table S2).

## Discussion

This is the first study from a randomised trial in Tanzania to examine age as a moderator of the effectiveness of a culturally adapted ED-based BI. Three main findings emerged. First, age groups moderated the effect of PPKAY on drinking days and number of drinks consumed but not on binge drinking. Second, in age-stratified analyses, young people showed intervention-specific reductions in drinking days and number of drinks, without corresponding reductions in binge drinking days or alcohol-related consequences. Third, reductions in alcohol use was observed over time across both age groups and study arms.

Age moderated the effect of PPKAY on drinking days and number of drinks consumed, with greater intervention-associated reductions observed among young people. While other ED-based studies have reported stronger effects of BIs among younger drinkers (19,23), meta-analytic evidence does not consistently demonstrate effect modification by age across settings (25) suggesting that such heterogeneity may be context- and outcome-specific. In the present study, moderation was evident for measures reflecting overall drinking patterns but not for binge drinking, indicating that age-related differences in intervention effect may depend on the behavioural domain assessed. Drinking frequency and total consumption capture broader patterns of alcohol use (33), which from a developmental perspective are more variable during young adulthood, a period characterised by role transitions and shifting trajectories of alcohol use over time (34,35). In contrast, alcohol use in adulthood may be more behaviourally entrenched and reinforced through habitual processes, contributing to the persistence of drinking patterns over time (35,36), which may attenuate intervention responsiveness among adults, contributing to greater reductions seen in young people. The absence of moderation for binge drinking may reflect differences in underlying behavioural mechanisms. Motivational interviewing primarily targets reflective processes (e.g., motivation and intention) (37,38), while binge drinking may arise from within-episode disinhibition and impaired control after drinking has begun (39). As these processes may operate similarly across age groups, age-related differences in intervention effect may be less apparent for this outcome. These findings suggest that age-related heterogeneity in intervention effect may be outcome-specific.

In age-stratified analyses, young people receiving PPKAY demonstrated greater reductions in drinking days and overall drinking quantity, while no significant changes were observed in binge drinking days or alcohol-related consequences. This pattern is consistent with prior evidence from ED-based BIs for adolescents and young people, which indicates that while brief motivational interventions can produce modest reductions in alcohol consumption, effects on high-risk episodic drinking and alcohol-related harms are often inconsistent, particularly over short follow-up periods (25,40,41). Binge drinking among young people is strongly shaped by social and peer contexts, which may limit the responsiveness of episodic heavy drinking to one-time, individually focused interventions delivered in ED settings (42,43). As a result, BIs may preferentially influence more readily modifiable aspects of drinking behaviour, such as drinking frequency or typical quantity. The absence of short-term changes in alcohol-related consequences likely reflects both their relative rarity and their temporal relationship with alcohol use. Changes in consumption typically precede measurable changes in harms, making short follow-up periods sensitive to reductions in drinking than changes in alcohol-related consequences (41,44). The absence of change in PHQ-9 scores suggests that reductions in alcohol consumption did not result in worsening depressive symptoms. Taken together, these findings suggest that ED-based BIs may have greater effects on drinking quantity and frequency than on binge drinking for young people, reflecting differences in behavioural processes underlying these drinking patterns.

Alcohol use declined over time across participants in both the PPKAY and usual care arms, across age groups. Similar trends have been reported in ED-based intervention trials, where reductions in drinking frequently occur irrespective of intervention assignment following injury-related presentations (17,45). This phenomenon is consistent with the sentinel event hypothesis, which proposes that alcohol-related injuries and other adverse health events can initiate self-directed behaviour change through heightened risk appraisal and self-reflection. even in the absence of structured intervention (46). From a life-course perspective, reductions in alcohol use may also reflect broader age-related changes in drinking behaviour.

Epidemiological evidence indicates that alcohol consumption and heavy drinking often decline beyond early adulthood as individuals transition into more stable social roles and responsibilities, a process commonly referred to as “maturing out” (47). Accordingly, these findings should not be interpreted as suggesting that ED-based brief alcohol interventions are unnecessary; rather, injury-related ED encounters may independently drive reductions in alcohol use, with additional intervention effects observed for some outcomes.

## Limitations

These findings should be interpreted in light of several limitations. This analysis examined age as a moderator within a pragmatic randomised trial, which was not specifically powered to detect subgroup effects; consequently, estimates of age-related interactions should be interpreted with caution, particularly among younger participants who comprised a smaller proportion of the sample. However, sensitivity analyses using age as a continuous variable yielded consistent patterns of findings, supporting the robustness of the observed results.

Alcohol use outcomes were based on self-report and may be subject to recall or social desirability bias, which could vary by age group or over time, although standardised instruments were applied consistently across study arms. In addition, we utilised a three-month follow-up time period, which may not have been sufficient to capture longer-term changes in alcohol-related consequences, as reductions in consumption often precede measurable changes in downstream harms. Despite these limitations, this analysis provides important insight into age-related heterogeneity in response to brief alcohol interventions in trauma care settings in LMICs.

## Implications

These findings have implications for clinical practice, public health, and future research in LMIC emergency care settings. Clinically, the results support the continued integration of brief, nurse-delivered alcohol interventions within EDs for injured patients with harmful or hazardous alcohol use, while underscoring the importance of age-responsive intervention approaches. Among young people, brief intervention was associated with greater reductions in drinking frequency and quantity, suggesting that these dimensions of alcohol use represent modifiable targets at this developmental stage. However, the absence of short-term effects on binge drinking and alcohol-related consequences suggest intervention effects may vary across outcomes. From a public health perspective, these findings support the integration of harm reduction approaches within trauma care systems in settings with a high burden of injury and risky alcohol use. Reductions observed across both intervention and usual care arms suggest that injury-related ED encounters may represent a window for behaviour change across age groups, within which BIs may provide additional benefit, particularly among young people. Future research should examine longer follow-up periods to assess whether reductions in alcohol use are sustained and lead to change in alcohol-related consequences and to investigate the behavioural and contextual mechanisms underlying the observed differences in intervention effects across outcomes and age groups.

## Conclusion

In this secondary analysis of a pragmatic ED-based randomised trial in Northern Tanzania, age moderated the effects of a culturally adapted BI on drinking days and number of drinks but not on binge drinking or alcohol-related consequences. Reductions in alcohol use were observed across both intervention and usual care groups in both age categories; however, additional intervention-associated reductions in drinking days and number of drinks were evident among young people aged 18–24 years. These findings indicate that ED-based brief alcohol interventions are outcome-specific and vary by age groups.

## List of Abbreviations

AUDIT: Alcohol Use Disorders Identification Test
BI: Brief Intervention
DiD: Difference in Difference
DrInC: Drinker Inventory of Consequences
ED: Emergency Department
IRR: Incidence Rate Ratio
KCMC: Kilimanjaro Christian Medical Centre
LMICs: Low- and Middle-Income Countries
MI: Motivational Interviewing
PHQ-9: Patient Health Questionnaire-9
PPKAY: Punguza Pombe kwa Afya Yako
PRACT: Pragmatic Randomized Adaptive Clinical Trial
SBIRT: Screening, Brief Intervention, and Referral to Treatment
SD: Standard Deviation
SSA: Sub-Saharan Africa
UC: Usual Care
WHO: World Health Organization

## Declarations

### Ethics approval and Consent to participate

Ethical approval for this secondary analysis was obtained from the KCMC University Ethics Committee (Certificate #2457) and the Tanzanian National Institute for Medical Research (NIMR/HQ/R.8a/Vol.IX/3425). Consent to participate was not required for this analysis, as it involved secondary use of de-identified data.

### Data Availability

The data analyzed in this study derive from the Punguza Pombe kwa Afya Yako (PPKAY) pragmatic randomized adaptive clinical trial and are available through the National Institute on Alcohol Abuse and Alcoholism Data Archive (NIAAADA), which is hosted within the National Institute of Mental Health Data Archive (NDA). Due to ethical and regulatory requirements, access to these de-identified participant-level data is restricted. Investigators seeking access must be affiliated with an NIH-recognized research institution, hold an active Federalwide Assurance, and submit a Data Use Certification for approval by the NDA Data Access Committee. The dataset is archived under Collection ID 3425 (Pro00103724). The analytic code used for this secondary analysis is available from the authors upon request.

### Author Contribution

W.C.M., C.A.S., J.R.N.V., B.T.M., S.E.M., and F.J.M. contributed to the conceptualisation of this study. W.C.M. conducted the statistical analyses and drafted the original manuscript. Methodological input regarding study design and analytic approach was provided by C.A.S., J.R.N.V., B.T.M., S.E.M., and F.J.M., while statistical guidance and review of analytic models were provided by J.R.N.V., B.M., J.V.P.S., and A.J.P. Interpretation of the findings was undertaken by C.A.S., J.R.N.V., B.T.M., S.E.M., F.J.M and K.M. Supervision was provided by C.A.S., J.R.N.V., B.T.M., S.E.M., and F.J.M. All authors critically revised the manuscript for important intellectual content and approved the final version.

## Acknowledgement

We thank the patients and staff of the Emergency Department at KCMC in Moshi, Tanzania, for their participation and support. We also thank the Global Emergency Medicine Innovation and Implementation (GEMINI) Research Center and the KCMC University, SoPH, for providing a collaborative academic environment and valuable scientific input through presentations and discussions that informed the interpretation of this secondary analysis.

