## Supplemental Table 1 for "Age as a moderator of a brief alcohol intervention among injury patients in Northern Tanzania"

**SUPPLEMENTARY**

**Table S1**: Age-stratified predicted means at baseline and 3 months, and mean predicted differences (95% CI) for study outcomes.

| **Outcome** | **Arm** | **Predicted mean Baseline** | **Predicted mean 3 months** | **Within-arm change (95% CI)** | **DiD (95% CI)^a^** | **p-value** |
| --- | --- | --- | --- | --- | --- | --- |
| **AGE 18–24 YEARS** |  |  |  |  |  |  |
| **Binge drinking days** | Usual care | 1.6 | 0.7 | −0.9 (−2.5, 0.8) |  |  |
|  | Intervention | 2.6 | 0.3 | −2.2 (−3.3, −1.2) | −1.4 (−3.3, 0.6) | 0.17^b^ |
| **Drinking days** | Usual care | 4.6 | 3.2 | −1.4 (−4.2, 1.4) |  |  |
|  | Intervention | 6.2 | 0.8 | −5.4 (−6.6, −4.1) | −4.0 (−7.1, −0.8) | 0.013^b^ |
| **Number of drinks** | Usual care | 16.4 | 12.7 | −3.7 (−14.9, 7.6) |  |  |
|  | Intervention | 35.5 | 4.4 | −31.1 (−41.3, −20.9) | −27.5 (−42.8, −12.2) | <0.001^b^ |
| **AUDIT score** | Usual care | 13 | 4.4 | −8.6 (−11.0, −6.2) |  |  |
|  | Intervention | 13.1 | 4.5 | −8.6 (−10.0, −7.2) | 0.0 (−2.8, 2.8) | 0.986^c^ |
| **DrInC score** | Usual care | 12.9 | 4.2 | −8.7 (−16.2, −1.2) |  |  |
|  | Intervention | 15.8 | 5.7 | −10.1 (−14.7, −5.5) | −1.4 (−10.2, 7.4) | 0.758^c^ |
| **PHQ-9 score** | Usual care | 3.8 | 2.9 | −0.9 (−2.5, 0.7) |  |  |
|  | Intervention | 3.7 | 3.6 | −0.1 (−1.0, 0.9) | 0.9 (−1.0, 2.7) | 0.366^c^ |
| **AGE ≥25 YEARS** |  |  |  |  |  |  |
| **Binge drinking days** | Usual care | 4 | 0.9 | −3.1 (−5.4, −0.8) |  |  |
|  | Intervention | 5 | 0.6 | −4.4 (−6.0, −2.8) | −1.3 (−3.8, 1.2) | 0.31^b^ |
| **Drinking days** | Usual care | 8 | 2.1 | −5.9 (−7.7, −4.0) |  |  |
|  | Intervention | 8.5 | 1.6 | −7.0 (−7.9, −6.0) | −1.1 (−3.2, 0.9) | 0.291^b^ |
| **Number of drinks** | Usual care | 42 | 9.1 | −32.9 (−45.5, −20.2) |  |  |
|  | Intervention | 45.2 | 9.1 | −36.0 (−43.0, −29.1) | −3.2 (−17.1, 10.8) | 0.654^b^ |
| **AUDIT score** | Usual care | 12.8 | 3.6 | −9.1 (−10.2, −8.0) |  |  |
|  | Intervention | 12.9 | 4.1 | −8.8 (−9.4, −8.1) | 0.4 (−0.9, 1.7) | 0.549^c^ |
| **DrInC score** | Usual care | 13.5 | 2.9 | −10.6 (−13.9, −7.3) |  |  |
|  | Intervention | 13.9 | 2.6 | −11.3 (−13.2, −9.3) | −0.6 (−4.5, 3.2) | 0.741^c^ |
| **PHQ-9 score** | Usual care | 2.9 | 3.9 | +1.0 (0.1, 1.9) |  |  |
|  | Intervention | 3.2 | 3.6 | +0.5 (−0.1, 1.0) | −0.5 (−1.5, 0.5) | 0.334^c^ |

Note:

^a^ Positive and negative values reflect the reduction or increase from predicted baseline values to 3-month follow-up, respectively.

^b^ Count outcomes (binge drinking days, drinking days, and number of drinks) were analysed using mixed-effects negative binomial models.

^c^ Continuous outcomes (AUDIT, DrInC, and PHQ-9 scores) were analysed using linear mixed-effects models.

Models were age-stratified and adjusted for sex and baseline AUDIT and PHQ-9 scores, with clustering at the practice level.

*p*-values represent the statistical significance of the difference-in-differences estimates
