## Supplemental Table 2 for "Age as a moderator of a brief alcohol intervention among injury patients in Northern Tanzania"

**Table S2**. Multiple imputation analysis: moderating effect of age on PPKAY intervention outcomes at 3 months

| **Outcome** | **Intervention effect (PPKAY vs Usual Care)** | **Age main effect (per +1 year)** | **Time × Intervention × Age (interaction)** | **p-value (interaction)** |
| --- | --- | --- | --- | --- |
| **Binge Drinking Days** | IRR = 0.74 (95% CI 0.24–2.26) | IRR = 1.04 (95% CI 1.00–1.08) | IRR = 1.02 (95% CI 0.94–1.11) | 0.663 |
| **Drinking Days** | IRR = 0.62 (95% CI 0.36–1.07) | IRR = 1.02 (95% CI 1.01–1.04) | IRR = 1.02 (95% CI 0.98–1.06) | 0.305 |
| **Number of Drinks** | IRR = 0.83 (95% CI 0.42–1.62) | IRR = 1.03 (95% CI 1.01–1.05) | IRR = 1.03 (95% CI 0.98–1.07) | 0.24 |
| **DrInC Score (log)** | β = 0.02 (95% CI −0.23 to 0.28) | β = −0.01 (95% CI −0.02 to 0.00) | β = −0.01 (95% CI −0.04 to 0.01) | 0.307 |
| **AUDIT Score (log)** | β = 0.10 (95% CI −0.07 to 0.27) | β = −0.01 (95% CI −0.02 to −0.00) | β = −0.01 (95% CI −0.02 to 0.01) | 0.503 |
| **PHQ-9 Score (log)** | β = 0.08 (95% CI −0.09 to 0.25) | β = 0.00 (95% CI −0.01 to 0.01) | β = 0.00 (95% CI −0.02 to 0.02) | 0.972 |
