## Supplemental Table 3 for "Age as a moderator of a brief alcohol intervention among injury patients in Northern Tanzania"

**Table S3**. Multiple imputation analysis: moderating effect of age on PPKAY intervention outcomes at 3 months

| **Outcome** | **Intervention effect (PPKAY vs Usual Care)** | **Age main effect (18–24 vs ≥25)** | **Time × Intervention × Age (interaction)** | **p-value (interaction)** |
| --- | --- | --- | --- | --- |
| **Binge Drinking Days** | IRR = 0.76 (95% CI 0.26–2.21), p=0.611 | IRR = 0.77 (95% CI 0.16–3.77), p=0.748 | IRR = 0.66 (95% CI 0.04–11.77), p=0.777 | 0.777 |
| **Drinking Days** | IRR = 0.76 (95% CI 0.42–1.38), p=0.371 | IRR = 1.44 (95% CI 0.49–4.23), p=0.508 | IRR = 0.28 (95% CI 0.08–1.02), p=0.054 | 0.054 |
| **Number of Drinks** | IRR = 1.01 (95% CI 0.49–2.10), p=0.969 | IRR = 1.34 (95% CI 0.44–4.09), p=0.604 | IRR = 0.18 (95% CI 0.04–0.81), p=0.026 | 0.026 |
| **DrInC Score (log)** | β = 0.03 (95% CI −0.19 to 0.26), p=0.773 | β = 0.25 (95% CI −0.38 to 0.88), p=0.442 | β = −0.18 (95% CI −0.97 to 0.60), p=0.649 | 0.649 |
| **AUDIT Score (log)** | β = 0.14 (95% CI −0.05 to 0.33), p=0.155 | β = 0.32 (95% CI −0.01 to 0.66), p=0.058 | β = −0.38 (95% CI −0.84 to 0.08), p=0.108 | 0.108 |
| **PHQ-9 Score (log)** | β = 0.09 (95% CI −0.09 to 0.27), p=0.334 | β = −0.01 (95% CI −0.40 to 0.38), p=0.960 | β = 0.33 (95% CI −0.14 to 0.81), p=0.166 | 0.166 |

Note:

Estimates are pooled across 20 multiply imputed datasets. Models were adjusted for sex and included baseline and 3-month observations.
